# Investigation of the differential biology between benign and malignant renal masses using advanced magnetic resonance imaging techniques (IBM-Renal): a multi-arm, non-randomised feasibility study

**DOI:** 10.1101/2024.05.03.24306816

**Authors:** Ines Horvat-Menih, Mary McLean, Maria Jesus Zamora-Morales, Marta Wylot, Joshua Kaggie, Alixander S Khan, Andrew B Gill, Joao Duarte, Matthew J Locke, Iosif A Mendichovszky, Hao Li, Andrew N Priest, Anne Y Warren, Sarah J Welsh, James O Jones, James N Armitage, Thomas J Mitchell, Grant D Stewart, Ferdia A Gallagher

## Abstract

**Introduction:** Localised renal masses are an increasing burden on healthcare due to the rising number of cases. However, conventional imaging cannot reliably distinguish between benign and malignant renal masses, and renal mass biopsies are unable to characterise the entirety of the tumour due to sampling error, which may lead to delayed treatment or overtreatment. There is an unmet clinical need to develop novel imaging techniques to characterise renal masses more accurately. Renal tumours demonstrate characteristic metabolic reprogramming, and novel MRI methods have the potential to detect these metabolic perturbations which may therefore aid accurate characterisation. Here we present our study protocol for the Investigation of the differential biology of Benign and Malignant renal masses using advanced magnetic resonance imaging techniques (IBM-Renal).

**Methods and analysis:** IBM-Renal is a multi-arm, single-centre, non-randomised, feasibility study with the aim to provide preliminary evidence for the potential role of the novel MRI techniques to phenotype localised renal lesions. 30 patients with localised renal masses will be recruited to three imaging arms, with 10 patients in each: (1) hyperpolarised [1-^13^C]-pyruvate MRI (HP ^13^C-MRI), (2) deuterium metabolic imaging (DMI), and (3) sodium MRI (^23^Na-MRI). The diagnosis will be made on samples acquired at biopsy or at surgery. The primary objective is to investigate whether novel MRI techniques can identify the differences between benign and malignant tumours, while the secondary objectives aim to assess how complementary the techniques are, and if they provide additional information. Exploratory objective will be to link imaging findings with clinical data and molecular analyses for biological validation of the novel MRI techniques.

**Ethics and dissemination:** This study was ethically approved (UK REC HRA: 22/EE/0136; current protocol version 2.1 dated 11/08/2022). The plans for dissemination include presentations at conferences, publications in scientific journals, a doctoral thesis, and patient and public involvement.

**Registration details:** ClinicalTrials.gov: <u>NCT06016075</u>

**Strengths and limitations of this study:**

- IBM-renal is the first prospective study to investigate the role of deuterium metabolic imaging and sodium MRI for the characterisation of indeterminate renal masses.
- Combining different MRI techniques in the same patient will allow a direct comparison and determining whether they provide additional data.
- The clinical team is multidisciplinary, enabling a multimodal assessment of these renal masses, including clinical, imaging, pathology data.
- Limitations of the study include potential pathological undergrading of benign renal masses, as some of these diagnoses are based on a single biopsy.
- As a feasibility study, the sample size is small, but the primary outcomes can be used to inform a large-scale study.

## Introduction

### Clinical need in management of localised renal masses

The incidence of renal cancer has increased significantly during the last two decades, with 13,322 new cases diagnosed annually in the UK, corresponding to an age-standardised incidence of 10.2 per 100,000 (1). The majority of these tumours are renal cell carcinomas (RCCs), which exhibit a variable degree of aggressiveness depending on the histology (2). Management options range from active surveillance through to radical surgical resection depending on the tumour type (3). An important unmet clinical need is that conventional imaging methods cannot reliably distinguish aggressive RCC subtypes from benign renal masses (4). Although an invasive renal mass biopsy is a key tool for discriminating radiologically indeterminate renal lesions, it is subject to sampling error which is particularly problematic in heterogeneous lesions. Biopsies may also be clinically challenging to perform, and are non-diagnostic in up to 20% of cases, which can result in either unnecessary or delayed surgery (3,5–7). Importantly, increased detection and improved management have not resulted in decreased mortality, suggesting there is an overdiagnosis and potentially overtreatment of benign renal tumours (1,8). As RCC remains one of the most lethal urological malignancies (9), there is a pressing need for novel methods to identify and characterise renal masses more accurately (10).

### Metabolic changes in RCC

Renal cell tumours harbour significant metabolic perturbations and different histologic subtypes have distinct metabolic phenotypes. In clear cell renal cell carcinoma (ccRCC), the major genetic driver is the loss of the von Hippel-Lindau (*VHL*) tumour suppressor gene, which leads to accumulation of hypoxia-inducible factor 1α(HIF1α) with downstream transcriptional activation of pathways involved in glycolysis (11,12). Metabolomic analyses have confirmed increased lactate labelling after [U-^13^C]glucose infusion in ccRCC tumours compared to adjacent normal kidney with glycolysis increasing in a grade-dependent manner (13–15). On the other hand, renal oncocytomas, which are classified as benign renal masses, suppresses oxidative metabolism due to defective complex I within the mitochondrial electron transport chain (11,16–18). Therefore, it is not clear to what extent the ratio between glycolytic and oxidative metabolism varies between benign and aggressive renal neoplasms (17,19). Recently studies have also reported mitochondrial respiration defects in both renal oncocytoma and its malignant counterpart, chromophobe RCC (chRCC), with the potential for differentiation based on genomic, transcriptomic, and metabolic levels (16,17,20). The question to be addressed in this study is whether MRI can be used to differentiate between benign and malignant lesions such as oncocytoma, chRCC, and ccRCC, and therefore early detection of renal malignancies requiring intervention.

### Imaging metabolism

Building upon the evidence described above, we hypothesise that novel MRI imaging techniques can non-invasively characterise whole-tumour metabolism and its heterogeneity across renal tumours. The most common clinical tool for assessment of tumour metabolism uses a radiolabelled glucose analogue, ^18^F-FDG, in conjunction with positron emission tomography (^18^F-FDG-PET), but is limited by renal excretion of tracer and has failed to show success in characterising renal tumours (21,22). ^99m^Tc-sestamibi in conjunction with Single-Photon Emission Computed Tomography (SPECT) has been used to detect a variable degree of tracer uptake in renal oncocytoma compared to chRCC in a small number of patients, which has been attributed to the tracer accumulation in cells with high levels of mitochondria (23,24). However, both techniques expose patients to ionising radiation, have poor soft tissue contrast, and the inability to detect specific downstream metabolites or their metabolic compartmentalisation (25,26).

Our multidisciplinary group are developing novel non-radioactive and non-invasive clinical tools for imaging tumour metabolism. For example, hyperpolarised [1-^13^C]-pyruvate MRI (HP ^13^C-MRI), following injection of intravenous hyperpolarised ^13^C-pyruvate, can be used to simultaneously detect glycolytic metabolism in the cytosol and oxidative metabolism in the mitochondria in tissue where there is sufficient metabolism (27). Two recent papers have shown that ^13^C-lactate labelling can be used to distinguish high grade ccRCC from lower grade tumours, driven by the pyruvate transporter (MCT1) (28). Furthermore, a case of a renal oncocytoma displayed the lowest pyruvate-to-lactate conversion compared to a range of malignant masses, including ccRCCs, suggesting that this may be a tool for discriminating these lesions (28).

More recently, we have implemented deuterium (^2^H) metabolic imaging (DMI) at clinical field strength as an alternative method for non-invasive detection of metabolism using orally administered deuterated glucose (29). This method can also be used to detect cytosolic lactate formation and mitochondrial oxidative metabolism, and is complementary to the information provided by HP ^13^C-MRI (30,31). We have undertaken this in the brain at clinical field strength (3T) and will apply it to the kidney in this study using dedicated hardware to assess whether differential glucose metabolism can be used to differentiate benign from malignant lesions.

### Imaging cellularity and structure

To complement the metabolic measurements, we will evaluate non-invasive methods to distinguish benign from malignant lesions based on differences in cellularity on histology. ^23^Na-MRI is a complementary tool to probe tissue structure as a measure of the tissue sodium concentration which is a function of cellularity due to the concentration gradient across the cell membrane. The method can also be used to extract an intracellular-weighted component of the sodium pool as a measure of the transmembrane sodium gradient (32). In the presence of hypoxia, the sodium/potassium adenosine triphosphate pump (Na^+^/K^2+^-ATPase) may be inhibited, leading to alterations in the sodium concentration (32,33). We have previously shown how ^23^Na-MRI can be used to demonstrate the Na^+^ gradient across the corticomedullary axis in the normal kidney and how this can be used to assess dynamic changes in renal sodium (33). We have also recently developed a novel birdcage MRI coil system for high resolution ^23^Na-MRI of the normal kidneys (34), which we will apply to imaging focal renal pathology as part of this study. We have shown the potential of ^23^Na-MRI in several cancer types, correlating with cellularity on histology, and will apply this to small renal masses for the first time here (35,36).

In addition, we have developed complementary ^1^H-MRI methods to quantitatively map T2 relaxation properties within tissue for assessment of diffuse renal disease (37). Here we will assess whether these can distinguish benign from malignant lesions. These parameters are dependent on the local tissue chemical properties and reflect the microenvironmental differences between benign and malignant disease.

### Rationale for the study

This pilot project will explore the role of multimodal MRI in characterising localised renal masses and how it can exploit the known biological differences between benign and malignant lesions. The project will assess if MRI can probe structure, function, and metabolism within the tumour and its microenvironment using three techniques:

1. HP ^13^C-MRI as a non-invasive measure of tissue metabolism following injection of hyperpolarised ^13^C-pyruvate to probe tumour lactate labelling.
2. DMI as an alternative method to probe both glycolytic and oxidative metabolism following oral deuterium labelled glucose to detect both lactate and the combined signal from glutamine+glutamate (Glx).
3. Measures of the tumour cellularity, heterogeneity, and membrane ion gradients using ^23^Na-MRI and fast, high-resolution measures of T2 relaxation.

The aim is to provide preliminary evidence for the potential role of these techniques to phenotype localised renal lesions and how they can be used as part of a larger multicentre study. The ultimate goal is to provide non-invasive tools for early detection of small aggressive renal tumours to enable timely surgical intervention.

## Methods and analysis

The study is reported in accordance to Standard Protocol Items: Recommendations for Interventional Trials (SPIRIT) checklist (38).

### Study design and objectives

The IBM-Renal (**I**nvestigation of differential biology of **B**enign and **M**alignant **Renal** masses using advanced magnetic resonance imaging techniques) study is designed as a feasibility study to acquire preliminary data to optimise future imaging protocols, and will be conducted as a non-randomised, physiological imaging study in patients with localised renal masses. The study will be conducted at a single site: Addenbrooke’s Hospital, Cambridge University Hospitals NHS Foundation Trust. For all imaging studies, the primary objective will be to investigate whether the methods can identify the differences between benign and malignant tumours, including measuring the ^13^C-pyruvate-to-lactate conversion with HP ^13^C-MRI, quantifying sodium concentration using ^23^Na-MRI, detecting ^2^H-glucose and its metabolites using DMI. Secondary objectives will aim to compare the imaging techniques to understand if they can produce complementary information. As an exploratory part of the study, the objective will be to link the imaging data with clinical data and tissue molecular analyses for biological validation of the novel MRI techniques.

### Participant selection

Participants will be identified through multidisciplinary team meetings or by clinical teams involved in their routine care at Addenbrooke’s Hospital, and recruited if they meet all the inclusion and none of the exclusion criteria as detailed in Table 1. The participants will be allocated into three imaging arms with 10 patients in each: (1) HP ^13^C-MRI; (2) DMI; and (3) ^23^Na-MRI. ^1^H-MRI with T2-mapping will be performed in all patients. The aim is to recruit at least four patients in each arm with an oncocytic renal neoplasm (mostly oncocytomas) and at least four patients with RCCs to enable direct comparison. Half of the patients in each arm will be selected from a cohort of newly diagnosed renal masses and imaged prior to biopsy, with 75-80% of these expected to have RCCs, and most of the remainder having oncocytic neoplasms. The other half will be acquired from retrospective cohorts of patients with previously diagnosed oncocytic neoplasms on active surveillance at least 6 weeks post biopsy. Diagnosis will be made on tissue samples acquired at biopsy or at surgery if applicable, using molecular markers where possible. This approach will ensure an appropriate balance between benign and malignant lesions in each cohort.

**Table 1:**
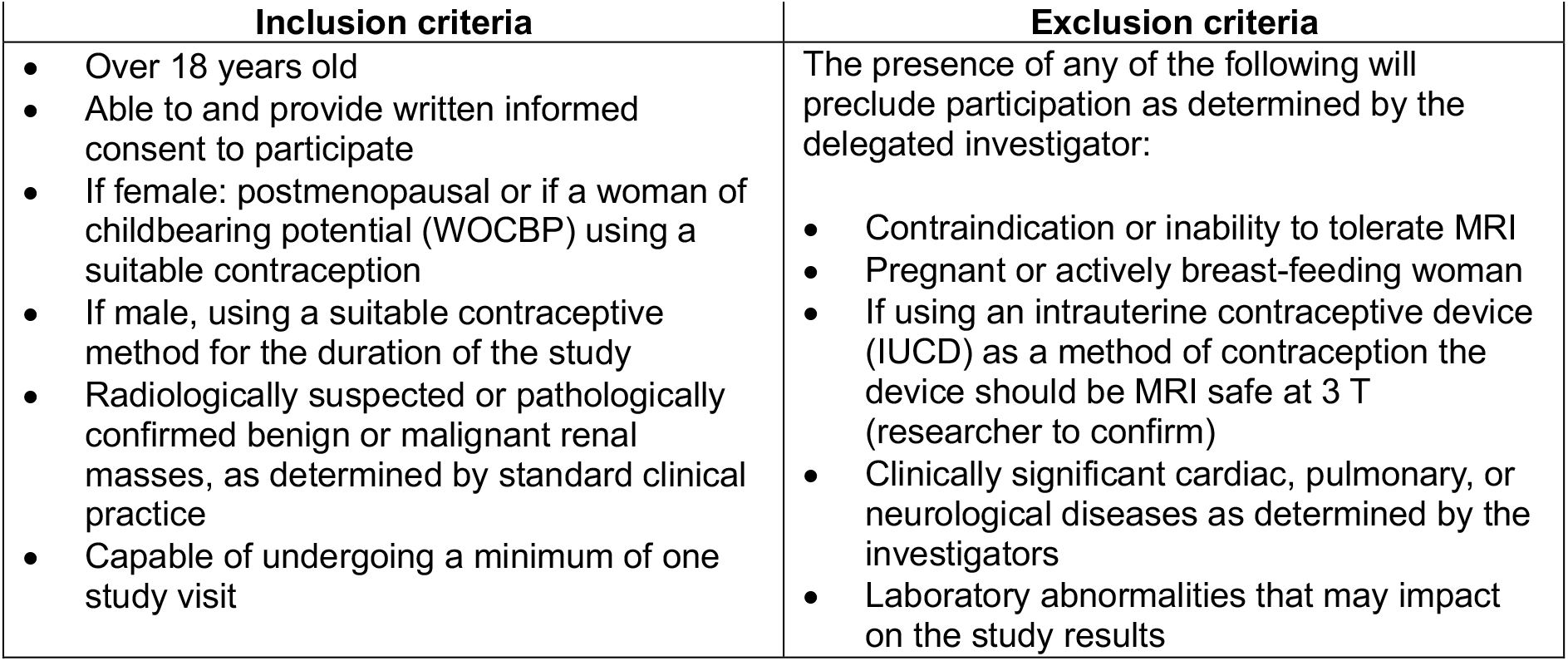
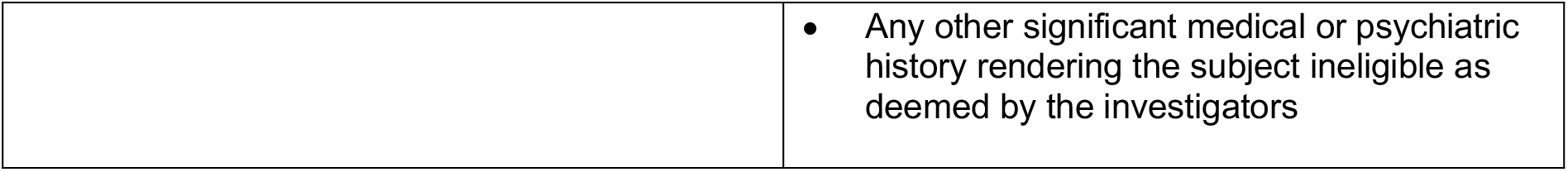
Inclusion and exclusion criteria for selection of study participants.

| Inclusion criteria | Exclusion criteria |
| --- | --- |
| • Over 18 years old<br>• Able to and provide written informed consent to participate<br>• If female: postmenopausal or if a woman of childbearing potential (WOCBP) using a suitable contraception<br>• If male, using a suitable contraceptive method for the duration of the study<br>• Radiologically suspected or pathologically confirmed benign or malignant renal masses, as determined by standard clinical practice<br>• Capable of undergoing a minimum of one study visit | The presence of any of the following will preclude participation as determined by the delegated investigator:<br>• Contraindication or inability to tolerate MRI<br>• Pregnant or actively breast-feeding woman<br>• If using an intrauterine contraceptive device (IUCD) as a method of contraception the device should be MRI safe at 3 T (researcher to confirm)<br>• Clinically significant cardiac, pulmonary, or neurological diseases as determined by the investigators<br>• Laboratory abnormalities that may impact on the study results |
|  | Any other significant medical or psychiatric history rendering the subject ineligible as deemed by the investigators |

Participants may be removed from the study at their choice or at the Investigator’s discretion if it is felt to be clinically appropriate. Reasons for participant withdrawal will be recorded. Primary reasons for withdrawal may include Serious Adverse Event (SAE), withdrawal of consent, lost to follow up, participant non-compliance, or study closed or terminated. Participants who are withdrawn from the study or do not complete at least one scan will be replaced.

### Interventions

Study participants will be deemed evaluable if they receive at least one scan on any of the three imaging techniques. Each study participant will be allocated a unique study number following study enrolment and will be identified by this number throughout the data collection and analysis process.

The participants will be asked to attend all or some of these timepoints:

1. Baseline imaging visit.
2. An optional repeat scan within seven days of the first scan using the same imaging technique.
3. For those not taking part in Part 2 above, an optional scan with another imaging technique within 14 days of the first scan.
4. An optional research biopsy will be undertaken at standard of care surgery for participants with a malignant renal mass, or at biopsy for participants with a benign lesion.

The study flow chart is presented in Figure *1*.

**Figure 1.**
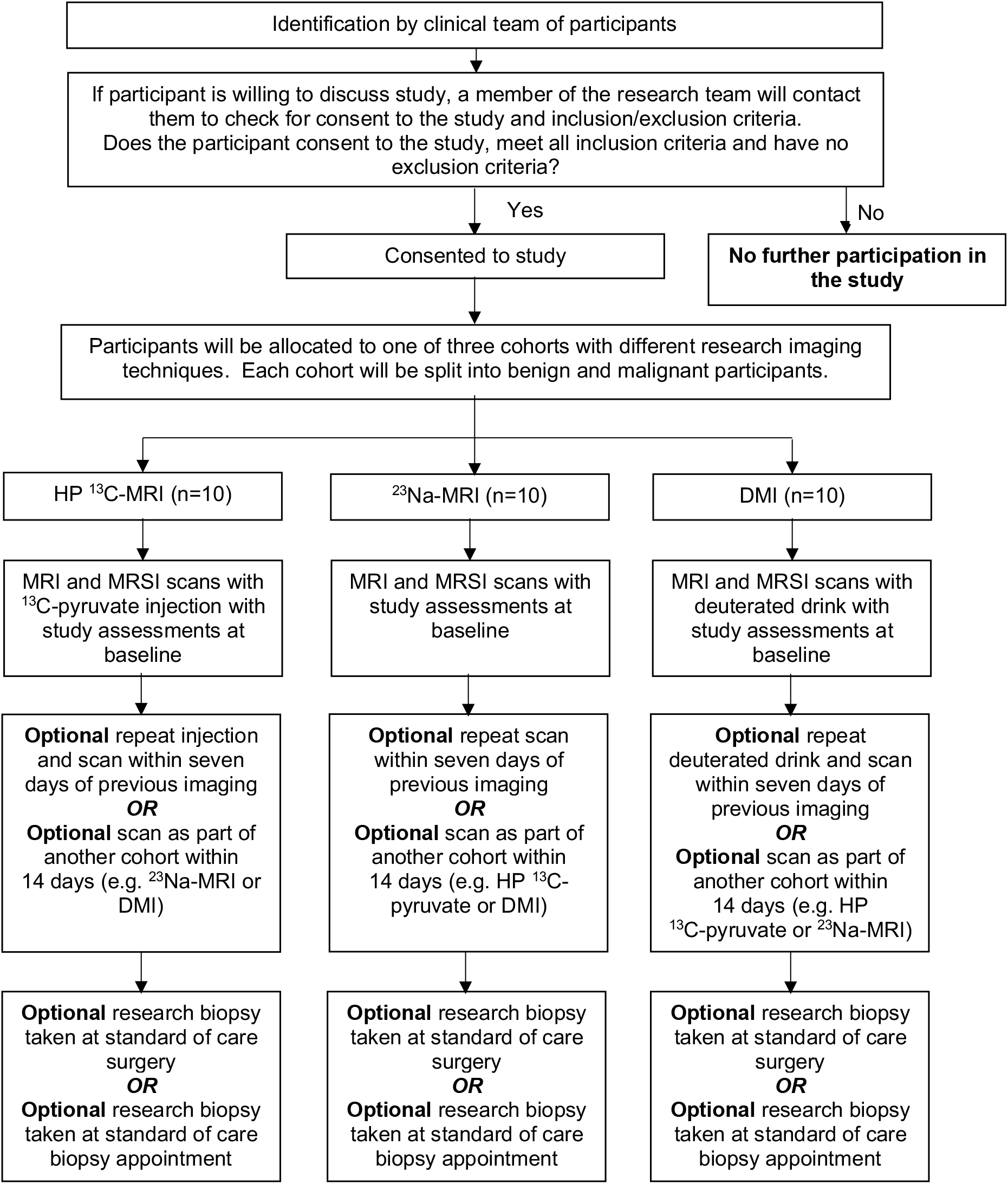
Study flow chart. MRSI = magnetic resonance spectroscopy imaging.

Table 2 provides an overview of assessments to be performed at each study visit.

**Table 2:** Schedule of assessments.

| Assessment | Visit/Days |  |  |  |
| --- | --- | --- | --- | --- |
|  | Screening <sup>a</sup> | Baseline MRI visit | Optional repeat MRI visit within seven days of baseline (if applicable) | Optional MRI visit with different imaging technique within 14 days of baseline (if applicable) |
| Attend Unit | * | * | * | * |
| Consent <sup>b</sup> | * | * | * | * |
| Medical History | * |  |  |  |
| Demography (weight, height, sex, Date of Birth) | * |  |  |  |
| Clinical Examination | * |  |  |  |
| ECOG performance score | * | * if clinically indicated | * if clinically indicated | * if clinically indicated |
| Venous blood sample <sup>c, d</sup> | * | * | * | * |
| Pregnancy test in WOCBP | * | * | * | * |
| MRI scan |  | * | * | * |
| General/additional assessments |  | * if clinically indicated | * if clinically indicated | * if clinically indicated |
| Vital Signs | * |  |  |  |
| • Pre-imaging |  | * | * | * |
| • Post-imaging |  | * | * | * |
| Injection of <sup>13</sup>C-pyruvate and/or deuterated glucose drink (for <sup>13</sup>C-pyruvate and DMI techniques only) |  | * | * | * |

\* Applicable assessment
- We will attempt to screen for participants during their standard of care visits and use of medical records. - Ongoing consent will be confirmed at each visit. - Venous blood samples will include but may not be limited to; full blood count, biochemical series and liver function test. - Full blood count samples will only be required at injection timepoints if they have not been taken recently i.e. within 14 days. These results will be copied in the study data.

For participants with benign renal masses that subsequently undergo active surveillance and have repeat MRI scans, we will seek the permission from the participants to review the clinically required MRI scan and compare to what was collected at the research scan. If the participant is not due to have a clinically required MRI scan, the research team will not affect this decision.

We will endeavour to minimise the number of injections and/or deuterated drinks for each participant, in as, the maximum number of injections/drinks will be limited to two each. Routes of administration for each of the imaging agents are as following:

- HP ^13^C-MRI: Single intravenous injection of up to 40 mL at 0.4 mL/kg of ^13^C-pyruvate through an intravenous cannula while the study participant is in position on the scanner bed. There will be an optional repeat injection of ^13^C-pyruvate at all imaging visits to test for repeatability of the ^13^C-MRI technique. These will take place within seven days of the first scan.
- ^23^Na-MRI: Not applicable. No additional research probe will be given to the participant as part of this imaging technique.
- DMI: At each imaging visit the participant will receive a deuterated glucose solution where 60 g of glucose is dissolved in 200 mL of water for injection (WFI) and the dose solution will be adjusted to their body weight at 0.75 g/kg body weight. There will be an optional repeat deuterated drink at all imaging visits to test for repeatability of the DMI technique. These will take place within seven days of the first scan.

### The sample size calculation and outcomes

The study has been powered to assess changes in the ^13^C-pyruvate metabolism from the data we collected from nine treatment-naïve renal tumour patients (28). This work showed that the median pyruvate-to-lactate conversion constant (*k*PL) in ccRCCs was 0.0065 (range 0.0024-0.0151), while it was 0.0043 (range 0.0028-0.0076) in the normal kidney. This study also reported metabolism in a single case of renal oncocytoma, which showed both the lowest conversion constant and lactate-to-pyruvate ratio. There are currently no published students assessing DMI and ^23^Na-MRI quantitative parameters in human kidney tumours.

Based on the parameters obtained from the HP-^13^C-MRI we therefore calculated the following sample sizes: we plan to include up to 30 participants in total: 15 with benign renal masses and 15 with malignant renal masses. These participants will be divided equally into three imaging arms (HP ^13^C-MRI, ^23^Na-MRI and DMI); therefore five benign and five malignant participants will be recruited to each imaging arm. If participants are willing to take part in the optional additional scan using a different imaging technique, these participants will be counted towards both arms of the study and therefore the total number recruited to the study will be less than 30 participants.

Descriptive statistics will be used. The primary covariates to be studied are as follows:

- HP-^13^C-MRI: ratio of the summed hyperpolarised ^13^C-lactate to the summed ^13^C-pyruvate over the timecourse of the experiment as a quantitative metric of pyruvate-to-lactate exchange catalysed by the enzyme lactate dehydrogenase (LDH). This metric is termed the lactate-to-pyruvate ratio (LAC/PYR). We have significant experience in developing quantitative methodology to analyse this data (39).
- ^23^Na-MRI: total sodium concentration (TSC), as a metric to quantify accumulation of Na+ in the tissue of interest. This metric was used in comparison between prostate cancer and normal prostate tissue(35).
- DMI: ratio of the summed ^2^H-lactate over the summed combined signal from ^2^H-glutamine+^2^H-glutamate (^2^H-Glx) as a measure of the ratio of glycolysis to oxidative metabolism (40).

### Data management and confidentiality

#### Case Report Form (CRF)

All data collected during the study will be collected or transferred into the CRF which will be anonymised. All study data in the CRF must be extracted from, and be consistent with, the relevant source documents. The CRFs must be completed, dated, and signed by the investigator or designee in a timely manner. The CRF will be accessible to relevant study team members, study monitors, auditors or inspectors as required.

#### Data protection and participant confidentiality

All investigators and study site staff involved in this study must comply with the requirements of the General Data Protection Regulation (GDPR) 2018 and Trust Policy with regards to the collection, storage, processing, and disclosure of personal information and will uphold core principles. The personal data recorded on all documents will be regarded as strictly confidential.

#### Study documentation and archiving

All essential source and study documentation including the Study Master File, source data, and proforma will be securely archived after the last analysis of the study data has been completed and the Final Study Report has been submitted to the relevant authorities. Archiving must be provided as per local policy or the length of time specified by current applicable legislation, whichever is the longer. The Investigator must not destroy any documents or records associated with the study without written approval from the Sponsor.

## Ethics and dissemination

### Ethical and Regulatory Considerations

Following the application through the Integrated Research Application System (IRAS, number: 314155), this study with related documentation has been approved by the East of England – Cambridge East Research Ethics Committee (REC), Health Research Authority (HRA), receiving the REC reference: 22/EE/0136. The Research & Development (R&D) Department of Cambridge University Hospitals NHS Foundation Trust and the University of Cambridge act as sponsors for this research project with respect to the UK Policy Framework for Health and Social Care Research. Further ethical and regulatory considerations are detailed below.

#### Informed Consent form

The Informed Consent form was approved by the REC and is in compliance with good clinical practice (GCP), local regulatory requirements, and legal requirements. The investigator must ensure that each study participant, or their legally acceptable representative, is fully informed about the nature and objectives of the study and possible risks associated with their participation. The suitably trained investigator will obtain written informed consent from each participant before any study-specific activity is performed. The investigator will retain the original of each signed informed consent form.

#### Research Ethics Committee review

Before the start of the study or implementation of any amendment we will obtain approval of the study protocol, protocol amendments, informed consent forms and other relevant documents e.g., advertisements and GP information letters if applicable from the REC. All correspondence with the REC will be retained in the Study Master File.

#### Regulatory issues

This study is not a Clinical Trial of an Investigational Medicinal Product (IMP) as defined by the European Union (EU) Directive 2001/20/EC and no submission to the Clinical Trials Unit at the Medicines and Healthcare products Regulatory Agency (MHRA) is required.

#### Protocol amendments

Protocol amendments must be reviewed and agreed by the Sponsor prior to submission to the REC/HRA. The only circumstance in which an amendment may be initiated prior to REC/HRA approval is where the change is necessary to eliminate apparent, immediate risks to the participants (Urgent Safety Measures). In this case, accrual of new participants will be halted until the REC/HRA approval has been obtained.

#### Declaration of Helsinki and Good Clinical Practice

The study will be performed in accordance with the spirit and the letter of the declaration of Helsinki, the conditions and principles of GCP, the protocol and applicable local regulatory requirements and laws.

#### GCP training

All study staff must hold evidence of appropriate GCP training or undergo GCP training prior to undertaking any responsibilities on this study. This training should be updated every 2 years or in accordance with Cambridge University Hospitals NHS Foundation Trust policy.

### Safety considerations

#### Adverse Reactions/ Expected Adverse Events

There are no expected adverse reactions (AR) associated with ^13^C-pyruvate and deuterated glucose MRI. If any ARs are observed during this study, they will be recorded on the proforma and reviewed by the research team.

The following adverse events (AE) are known side effects of the assessment procedures:

- Bruising at the sites of venepuncture.
- For those participants having the ^13^C-pyruvate injection, a transient local reaction at site of injection, a transient change in taste, and mild flushing.

They are generally not serious in nature and will not be recorded in the AE/AR log as part of this study.

Participants with solid malignancies are expected to have cancer and treatment related adverse events and some of them may be serious adverse events (SAE). However, as these are related to cancer rather than the study procedures they will not be recorded or collected as study data during this study. Only study procedure related SAE will be recorded.

#### Recording, evaluation and reporting of adverse events

The Sponsor expects that all adverse events are recorded from the point of Informed Consent. All AR/AEs will be assessed by the investigator and recorded in medical notes as well as on the proforma (except for expected AEs and SAEs related to cancer).

Individual adverse events should be evaluated by the Investigators. This includes the evaluation of its seriousness, causality, severity, and any relationship between the medicinal product(s) and/or concomitant therapy and the adverse event.

The Chief Investigator is responsible for the prompt notification to the Sponsor and the REC that gave a favourable opinion of the study where in the opinion of the Chief Investigator the event was:

- “related”: that is, it resulted from administration of any of the research procedures; and
- “unexpected”: that is, the nature and severity of the event is not listed in the protocol or the investigators brochure as an expected occurrence.

Reports of related and unexpected SAEs should be submitted to the Sponsor and the REC within 15 days of the Chief Investigator becoming aware of the event.

*Toxicity – Emergency Procedures*

No toxicity is expected as both pyruvate or glucose are endogenous products. However, in the event of an acute hypersensitivity reaction, supportive care will be given to the participant according to local clinical procedures.

#### Patient and Public Involvement (PPI)

Patient representative groups were closely involved in preparation of the Study protocol. We have sought help from the Cambridge University Hospitals PPI Panel, who kindly reviewed the study documentation we were intending to submit with the IRAS for the REC review. With their valuable feedback we have adapted documentation to make it easier to follow, such as developing graphic representations of the study procedures, and preparing separate documents for each of the imaging arms.

#### Dissemination plans

The clinical and feasibility data are expected to be of great interest to the uro-oncological community, including radiologists, pathologists, surgeons, and oncologists. Results will be reported internally, presented at conferences, published in peer reviewed scientific journals, and will constitute a part of a PhD thesis. Further, we will engage patients and the public by organising workshops reporting the findings of the study and presenting at the public engagement festivals.

## Data Availability

No data was generated for this article.

## Authors’ contributions

F.A.G. is the chief investigator and initiated the collaborative project. I.H.M. and M.J.L conceptualised the study, developed the study design, drafted, and revised the study protocol and documentation. I.H.M. and M.J.Z.M. monitored data collection, drafted the paper. M.W. is the study coordinator, provided management oversight and monitored data collection. J.K., A.S.K., J.D., A.B.G., A.N.P., H.L., and M.A.M. contributed to the study design and statistical analysis plan. I.A.M, A.Y.W., S.J.W., J.O.J., J.N.A., T.J.M., and G.D.S. provided clinical expertise for the study design. All authors revised the paper.

## Acknowledgements

We acknowledge the invaluable feedback by the patient representatives, as well as the administrative and technical support from the Advanced Cancer Imaging and Urological Malignancies programmes, Cancer Research UK (CRUK) Cambridge Centre, and radiographers of the Magnetic Resonance Spectroscopy Unit, Addenbrookes.

